# The Bilateral Remote Ischaemic Conditioning in Children (BRICC) trial: a two-centre, double-blind, randomised controlled trial in young children undergoing cardiac surgery

**DOI:** 10.1101/2023.04.21.23288646

**Authors:** Nigel E Drury, Carin van Doorn, Rebecca L Woolley, Rebecca J Amos-Hirst, Rehana Bi, Collette M Spencer, Kevin P Morris, James Montgomerie, John Stickley, Adrian Crucean, Alicia Gill, Matt Hill, Ralf J.M. Weber, Lukas Najdekr, Andris Jankevics, Andrew D. Southam, Gavin R. Lloyd, Osama Jaber, Imre Kassai, Guiseppe Pelella, Natasha E Khan, Phil Botha, David J Barron, Melanie Madhani, Warwick B Dunn, Natalie J Ives, Paulus Kirchhof, Timothy J Jones

## Abstract

**Background and Aims:** Previous trials evaluating remote ischaemic preconditioning in children undergoing cardiac surgery showed mixed results. We sought to determine whether adequately delivered bilateral preconditioning is cardioprotective in young children, with or without cyanosis, undergoing surgery.

**Methods:** Prospective, double-blind, randomised controlled trial at two UK centres. Children aged 3-36 months undergoing tetralogy of Fallot repair or ventricular septal defect closure were randomised in a 1:1 ratio to receive either bilateral preconditioning or sham intervention. Participants were followed up until hospital discharge or 30 days. The primary outcome was area under the curve for high-sensitivity troponin-T in the first 24 hours after surgery, analysed by intention-to-treat. Right atrial biopsies were obtained in selected patients. Trial registration: ISRCTN12923441.

**Results:** Between 24 October 2016 and 8 December 2020, 120 eligible children were randomised to receive either bilateral preconditioning (n=60) or sham intervention (n=60). Participants had a median age of 7 months and 42 (35%) were female. The primary outcome, area under the curve for hs-troponin-T was higher in the preconditioning group (mean: 70.0±50.9µg/L/hr, n=56) than in controls (mean: 55.6±30.1µg/L/hr, n=58), p=0.04. Sub-group analyses did not show a differential treatment effect by oxygen saturations (p_interaction_=0.25) but showed evidence of differential treatment effect by underlying defect (p_interaction_=0.04). Myocardial metabolism, quantified in atrial biopsies, and secondary outcomes were not different between randomised groups.

**Conclusions:** Bilateral remote ischemic preconditioning does not attenuate myocardial injury in children undergoing surgical repair for congenital heart defects, and there was evidence of potential harm in unstented tetralogy of Fallot.

## INTRODUCTION

Myocardial protection against ischaemic-reperfusion injury is a key determinant of heart function and outcome following cardiac surgery in children [1]. With current strategies, myocardial injury occurs routinely following aortic cross-clamping, as quantified by a rise in circulating troponin concentrations in the first 24 hours after surgery [2,3]. This myocardial damage frequently impairs ventricular function, which may manifest as low cardiac output and require inotropic support in the early postoperative period. This is a major cause of morbidity and death in the early postoperative period [1,4] and children with preoperative cyanosis may be more vulnerable to the effects of ischaemia-reperfusion than acyanotic children [5,6].

Remote ischaemic preconditioning (RIPC), the application of brief, non-lethal cycles of ischaemia and reperfusion to a distant organ or tissue, is a simple, low-risk and readily available technique which may improve myocardial protection. Previous trials of RIPC in children undergoing cardiac surgery have shown mixed results [7–15] and have been criticised for a potentially inadequate stimulus; a manual sphygmomanometer may allow sub-clinical reperfusion during the ischaemic phase of RIPC [13], and the use of propofol anaesthesia has been suggested to interfere with the preconditioning pathway [16,17]. In addition, they have not evaluated the effects of preoperative cyanosis on RIPC [18] and have only applied the cuff to a single limb, potentially delivering a subtherapeutic stimulus in young children with a lower skeletal muscle mass compared to adults.

To provide a robust answer to the question of whether RIPC can attenuate perioperative myocardial injury in young children undergoing cardiac surgery, we conducted a randomised, prospective, double-blind trial comparing state-of-the-art bilateral preconditioning to a sham control in children with the two most common congenital heart defects requiring surgery [19] and investigated the impact of RIPC on myocardial metabolism during cardioplegic arrest.

## METHODS

### Study design

The Bilateral Remote Ischaemic Conditioning in Children (BRICC) trial was a double-blind, prospective, parallel group, randomised controlled trial in young children undergoing elective cardiac surgery at two paediatric cardiac surgical centres in the United Kingdom, Birmingham Children’s Hospital and Leeds Children’s Hospital. The study was approved by the West Midlands-Solihull NHS Research Ethics Committee (16/WM/0309, 5 August 2016). A detailed description of the methods is contained in the published trial protocol [20] and Supplementary materials.

### Patients

All infants and young children, aged 3 months to 3 years at the time of surgery, undergoing either complete repair of tetralogy of Fallot (TOF) or surgical closure of an isolated ventricular septal defect (VSD), with or without concomitant atrial septal defect (ASD) closure or pulmonary artery repair/augmentation, were eligible. Children were excluded if they: required an additional procedure (other than ASD closure or pulmonary artery repair/augmentation); had significant airway or parenchymal lung disease, bleeding disorder or a recent ischaemic event; had undergone previous cardiac surgery with cardioplegic arrest; required emergency surgery; or their parents declined to give consent. Initially those with a known major chromosomal defect were also excluded, as in previous trials [7,13], but this was amended during the trial as it became clear that there was no biological reason for exclusion relating to RIPC. Eligible patients were identified from multi-disciplinary team meetings, clinics, or surgical waiting lists. A parent information sheet was provided, either in person or sent in the post, and written informed consent was obtained by a Consultant Surgeon prior to enrolment.

### Randomisation and blinding

Participants were randomised 1:1 to receive either RIPC or a sham procedure (control) by the research nurse using a secure online randomisation system, developed and maintained by Birmingham Clinical Trials Unit, with a minimisation algorithm incorporating the following factors: 1) congenital heart defect; 2) presence of a right ventricular outflow tract (RVOT) stent in those with tetralogy of Fallot [21], and 3) surgical centre.

The trial intervention was delivered by an independent healthcare professional, trained and competent in delivering the intervention according to a standard operating procedure, who also performed the randomisation and was not involved in postoperative care. Blinding was maintained by covering the child with a surgical drape from above the nipples downwards including all four limbs throughout the period of cuff application, intervention, and removal. The research nurse, surgical, anaesthetic, perfusion, and paediatric intensive care unit (PICU) teams involved in the child’s care were blinded to group allocation throughout the trial.

### Procedures

After induction of anaesthesia but prior to sternotomy, the treatment group received RIPC induced simultaneously in both lower limbs using the PTSii digital tourniquet system (Delfi Medical Innovations, Vancouver) inflated to at least 50mmHg above systolic pressure for three cycles of 5-minutes ischaemia and 5-minutes reperfusion [22]; if one lower limb was unavailable, the cuff was placed on the upper arm instead. Continual loss of arterial flow during ischaemia was confirmed by concealed distal pulse oximetry [11] and if the distal trace was not rapidly lost, the cuff was tightened, or the inflation pressure increased to achieve arterial occlusion. Once the intervention had begun, each cuff was kept on the same limb to ensure repeated doses of ischaemia-reperfusion to the same muscle mass. In the control group, the cuffs were applied to a plastic tubing dummy limb placed between the patient’s legs and three sham inflation-deflation cycles performed, covered by a drape before, during and after the sham intervention to maintain blinding. Adherence to intervention was defined as receiving the allocated intervention, with documented loss of arterial flow during each period of limb ischaemia in the RIPC group.

All other aspects of anaesthesia, surgery, perfusion, and postoperative care were at the discretion of the blinded clinical team, without influence from the researchers, except for propofol, which was not used for induction or maintenance of anaesthesia, with isoflurane as the preferred volatile anaesthetic agent [23]. St Thomas’ cardioplegia was used at both sites and delivered according to local practice. Myocardial reperfusion on first release of the aortic cross-clamp was considered as time zero for postoperative events. Blood was drawn prior to sternotomy and at 3, 6, 12 and 24 hours after reperfusion, and plasma high-sensitivity (hs) troponin-T concentrations were quantified in batches using the fifth-generation Elecsys Tn-T HS assay (Roche, Basel) at an approved core laboratory.

Right atrial myocardial samples were obtained soon after aortic cross-clamping (onset ischaemia) and just before its release (late ischaemia) for metabolic phenotyping. Briefly, tissue extracts were analysed using ultra high-performance liquid chromatography-mass spectrometry (UHPLC-MS). Two complementary assays were applied: HILIC assay to study water-soluble metabolites; and C_18_ reversed-phase assay to determine changes in lipids during ischaemia. The impact of RIPC on myocardial metabolism was assessed through robust statistical analysis using correction for multiple testing and pathway enrichment analysis; for a detailed description, see Supplementary materials.

### Outcomes

The primary outcome was area under the time-concentration curve (AUC) for plasma hs-troponin-T release in the first 24 hours after aortic cross-clamp release (reperfusion) as a biomarker of myocardial injury. Secondary outcomes were: peak hs-troponin-T in the first 12 hours; total vasoactive inotrope score in the first 12 hours [24,25]; arterial lactate and central venous oxygen saturations in the first 12 hours; length of postoperative stay in the PICU; and length of postoperative stay in the hospital. Cardiac index in the first 12 hours was measured using ICON (Osypka Medical, Berlin) as an exploratory outcome in Birmingham only (see Supplementary materials).

The following serious adverse events were reported to the sponsor within 48 hours of identification: death; extracorporeal life support (ECLS); major neurological event; and further surgery or catheter intervention in the early post-operative period. Follow-up was until discharge from hospital or 30 days, whichever was sooner.

### Sample size

It was hypothesised that RIPC would reduce the AUC for hs-troponin-T release in the first 24 hours compared with controls, but that exposure to chronic hypoxaemia may impact on this reduction. Based on limited published data using a standard (fourth-generation) troponin assay, the proposed sample size was sufficient to detect a 35% reduction in postoperative troponin release, assuming a mean release equivalent to 350 μg/L/h in the control group compared with 228 μg/L/h in the RIPC group (extrapolated from the similarly mixed cohort of cyanotic and acyanotic children [7]), with a variability of 220 μg/L/h [10]. A sample size of at least 52 children per treatment group was needed to have a power of 80% and a significance level of 0.05 (2-sided). We therefore aimed to recruit up to 120 children to allow for dropouts, randomised in a 1:1 ratio between RIPC and control.

### Statistical analysis

Primary analysis of the primary and secondary outcome measures was performed according to the intention-to-treat principle. Analyses were undertaken using SAS v9.4. The primary comparison was comparing the RIPC group with the control group and all estimates of differences are presented with 95% confidence intervals.

To calculate the primary outcome, AUC for hs-troponin-T release in the first 24 hours, data were collected at baseline (pre-sternotomy) and at 3, 6, 12 and 24 hours after aortic cross-clamp release. The AUC was calculated using the trapezoidal method and compared between the RIPC and control group using a linear regression model, adjusting for the minimisation variables (congenital heart defect and centre) and baseline troponin. Missing baseline troponin values were imputed using the median value of the participant’s randomised group and type of congenital heart defect, whilst any missing postoperative value led to exclusion from the primary analysis. Subgroup analyses were also carried out on the primary outcome to assess whether there was evidence that the treatment effect differs by: preoperative oxygen saturations (cyanotic <90% or acyanotic ≥90%); congenital heart defect (TOF with RVOT stent, TOF without RVOT stent, or VSD); and age (<1 year or ≥1 year).

For the secondary outcomes, continuous data items (eg. peak troponin) were also analysed using a linear regression model. Continuous outcomes measured across more than three time points (eg. arterial lactate and central venous oxygen saturations) were analysed using mixed effect repeated measures models. Time to event data outcomes were analysed using a Cox regression model. P-values are reported from two-sided tests at the 5% significance level. The statistical analysis plan was agreed and signed-off prior to data lock, and the Chief Investigator and trial statisticians had access to the final dataset.

The trial was prospectively registered (ISRCTN12923441) prior to recruiting the first patient. It was overseen by an independent Data Monitoring Committee (see acknowledgements), who met at regular intervals during recruitment to review efficacy and safety data, and audited by the independent Clinical Research Compliance team, University of Birmingham. The first author is Chief Investigator of the trial and takes responsibility for the integrity of this report. All authors have read and agree to the manuscript as written. The funder had no role in study design, data collection, analysis, or interpretation, or writing of the report.

## RESULTS

Between 24 October 2016 and 8 December 2020, 306 children were screened, of whom 223 (72.9%) met the patient eligibility criteria and of these, 82 (36.8%) were excluded for logistical or other reasons (figure 1); 20 (14.2%) parents of otherwise eligible children declined consent. Recruitment was paused between 13 March 2020 and 29 June 2020 due to the impact of the Covid-19 pandemic on the National Health Service (see Supplementary materials). One hundred twenty-one infants/young children were randomised: 61 to RIPC; and 60 to control. One child in the RIPC group did not proceed to surgery so was excluded from the analysis, leaving 60 participants in each group who underwent surgery and completed follow-up. Of these, complete postoperative troponin data was available on 114 children (56 RIPC, 58 control) who were included in the primary outcome analysis. Baseline characteristics were similar between groups (table 1) and operative data is shown in table 2. Adherence to treatment allocation was achieved in 116 (96.7%) patients, 56 (93%) in the RIPC group, who received three confirmed cycles of limb ischaemia-reperfusion, and 60 (100%) in the control group. There was one (<1%) protocol deviation with propofol inadvertently used for induction of anaesthesia in a patient in the RIPC arm.

**Figure 1.**
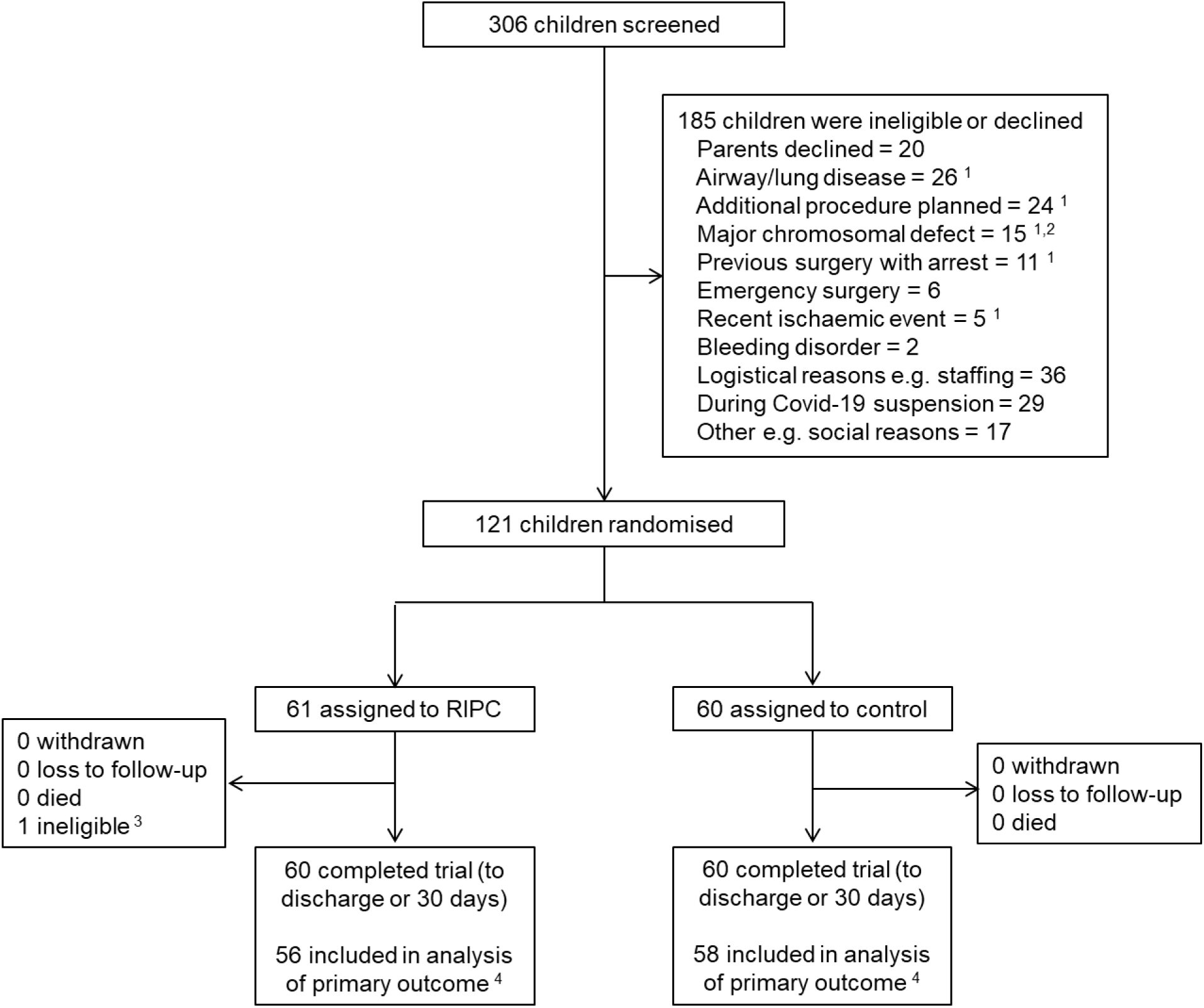
CONSORT flow diagram. ^1^ Patients counted in multiple categories due to concomitant reasons for exclusion: 191 reasons in 185 patients. ^2^ Prior to change to exclusion criteria to allow recruitment. ^3^ Participant was randomised in error, later found to be ineligible, did not undergo surgery and therefore outcome data was not available. Excluded from the trial post-randomisation, prior to the receipt of any intervention and not included in subsequent analyses. ^4^ With complete primary outcome data available. RIPC, remote ischaemic preconditioning.

**Table 1.**
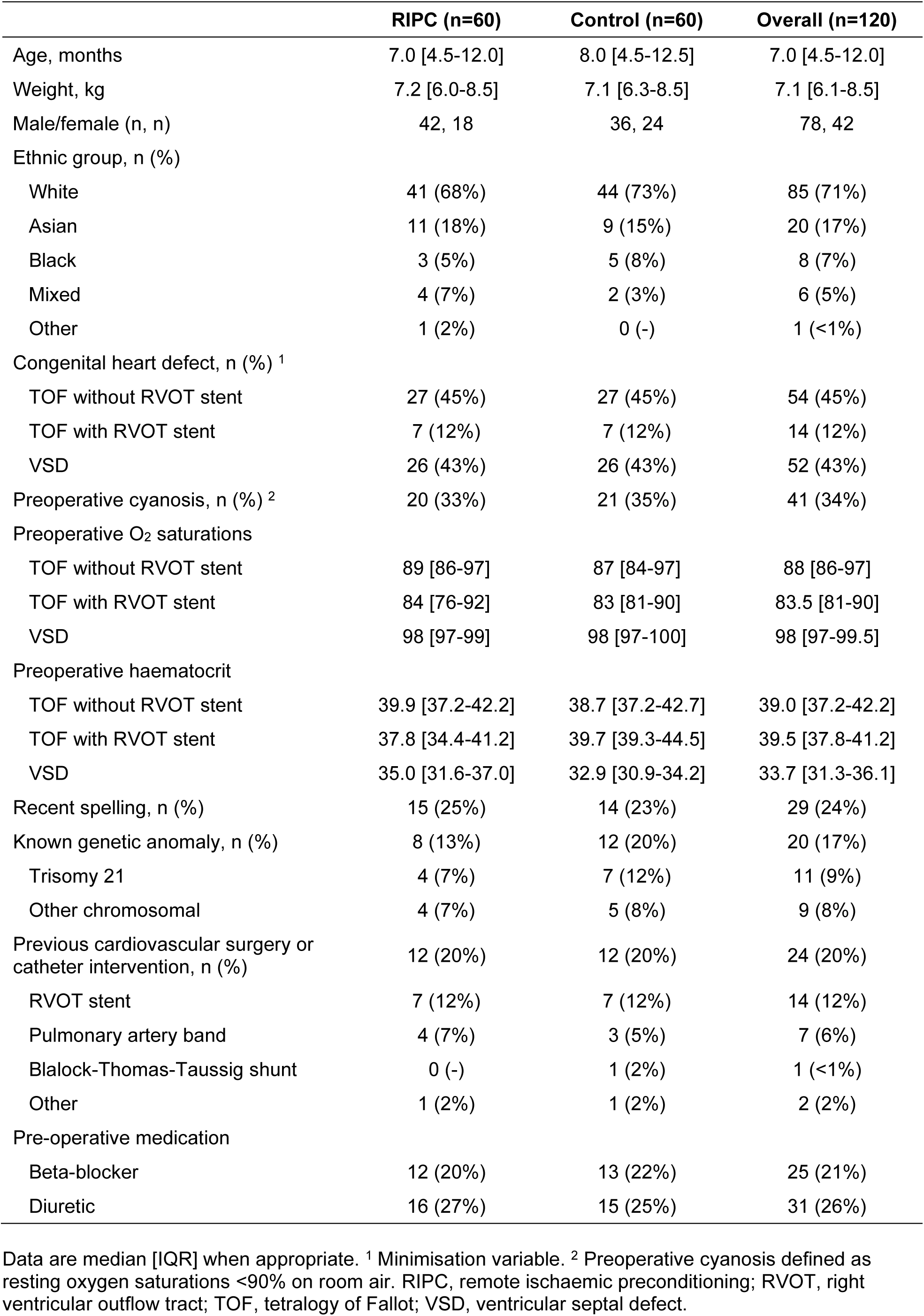
Baseline patient characteristics, by treatment group and overall.

**Table 2.** Operative data, by treatment group.

|  | **RIPC (n=60)** | **Control (n=60)** |
| --- | --- | --- |
| Anaesthesia |  |  |
| Volatile anaesthetic agent: isoflurane | 56 (93%) | 55 (92%) |
| Volatile anaesthetic agent: sevoflurane | 4 (7%) | 5 (8%) |
| MAC volatile anaesthetic pre-CPB, % | 0.7 [0.6-0.9] | 0.8 [0.7-1.0] |
| Time from start of intervention to aortic XC, minutes | 93 [81-105] | 92 [77-103] |
| In tetralogy of Fallot repair | (n=34) | (n=34) |
| RVOT muscle resected | 34 (100%) | 33 (97%) |
| RVOT stent removed <sup>1</sup> | 6 (86%) | 7 (100%) |
| RVOT/TA/PA patch used | 32 (94%) | 33 (97%) |
| RV-PA conduit used | 2 (6%) | 1 (3%) |
| In isolated VSD closure | (n=26) | (n=26) |
| RVOT muscle resected | 5 (19%) | 3 (12%) |
| VSD closed | 60 (100%) | 59 (98%) <sup>2</sup> |
| Total CPB time, minutes | 91.0 [67.0-114.0] | 87.5 [70.0-106.5] |
| Repeat CPB required | 7 (12%) | 5 (8%) |
| Total aortic XC time, minutes | 64.0 [45.0-84.5] | 58.5 [44.5-74.0] |
| Repeat aortic XC required | 6 (10%) | 5 (8%) |
| Type of cardioplegia used |  |  |
| Cold blood | 54 (90%) | 53 (88%) |
| Cold crystalloid | 6 (10%) | 6 (10%) |
| Warm blood | 0 | 1 (2%) |
| Number of cardioplegia doses | 2 [2-3] | 2 [2-3] |
| Total cardioplegia volume, ml | 360 [272-450] | 369 [294-450] |
| Post-operative recovery |  |  |
| Drain loss at 12 hours, ml | 55.0 [45.0-87.5] | 79.5 [40.0-107.5] |
| Blood transfusion post-CPB in first 12 hours | 13 (22%) | 11 (18%) |
| Volume blood transfused in first 12 hours, ml/kg in those transfused | 75 [55-100] | 50 [30-80] |
| Haemoglobin at 12 hours, g/dl | 119 [111-134] | 119 [107-130] |
| Time to extubation, hours | 6.3 [3.7-15.2] | 6.9 [3.6-13.3] |

Data are median [IQR] when appropriate. <sup>1</sup> Of the 7 patients in each treatment arm with tetralogy of Fallot and an RVOT stent. <sup>2</sup> Small infant with tetralogy of Fallot and bilateral superior vena cavae in whom an RV-PA conduit was placed but complete repair was abandoned. CPB, cardiopulmonary bypass; MAC, minimum alveolar concentration; PA, pulmonary artery; RIPC, remote ischaemic preconditioning; RVOT, right ventricular outflow tract; RV-PA, right ventricle to pulmonary artery; TA, transannular; VSD, ventricular septal defect; XC, cross-clamp.

### Primary outcome

Mean AUC for hs-troponin-T was 13.2µg/L/hr higher (i.e. worse) in the RIPC group when compared with control (Mean diff: 13.2; 95% CI: 0.5, 25.8; p=0.04) (figure 2 and table 3). The per-protocol analysis supported the primary analysis with mean AUC remaining higher in the RIPC group (Mean diff: 15.5; 95% CI: 2.6, 28.3; p=0.02). Prespecified sub-group analyses did not show a differential treatment effect in cyanotic and acyanotic children (interaction p-value = 0.25); however, there was evidence of an interaction by congenital heart defect (interaction p-value = 0.04), potentially driven by a greater AUC for troponin release in children with unstented TOF receiving RIPC, though numbers were small: unstented TOF, mean diff: 30.9, 95% CI: 12.2, 49.6; stented TOF, mean diff: 7.8, 95% CI: -27.7, 43.4; VSD, mean diff: -3.2, 95% CI: -22.0,15.5 (figure 3 and table 4). There was also some evidence of an interaction between age and RIPC (interaction p-value = 0.06), independent of type of defect.

**Figure 2.**
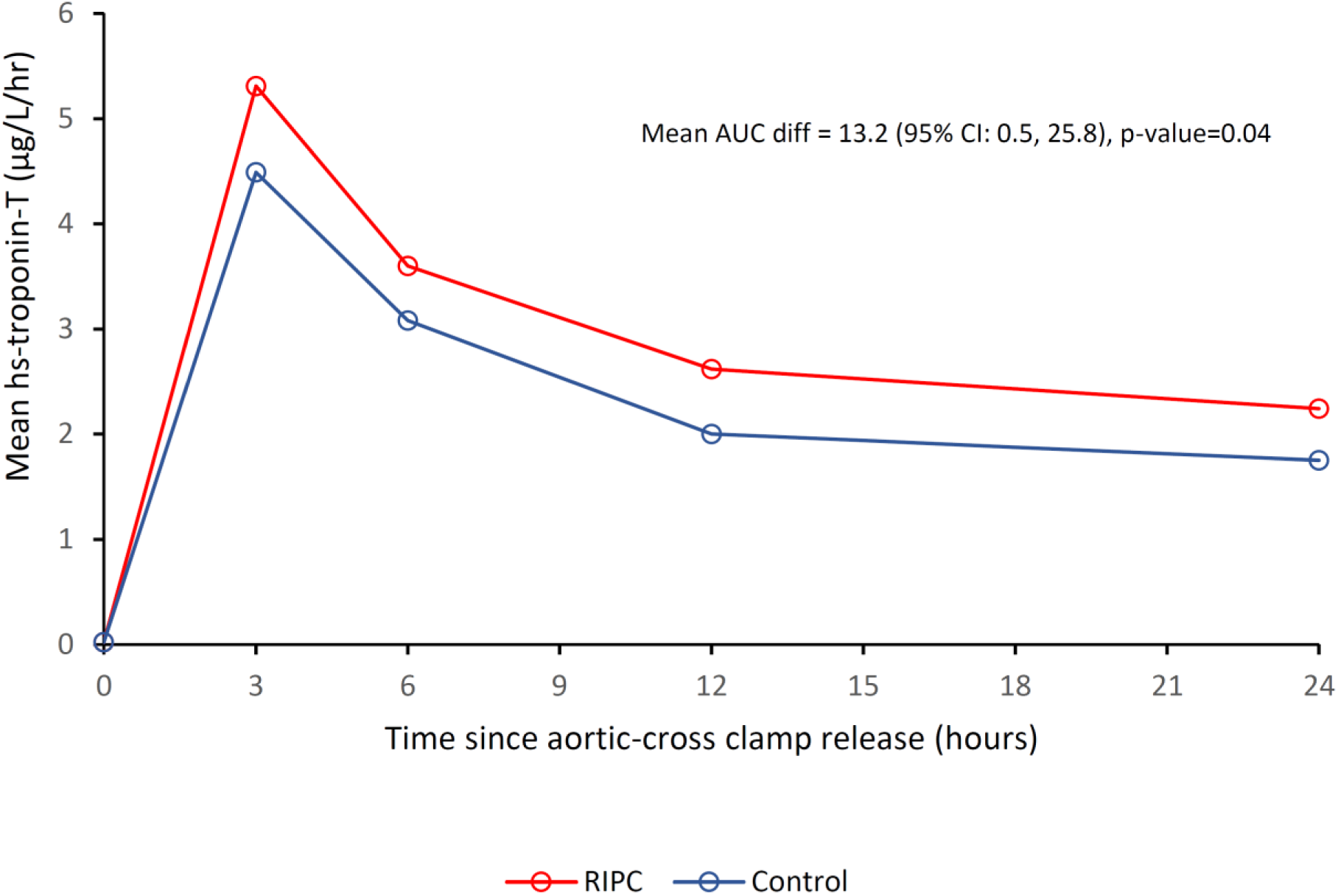
Mean hs-troponin-T release in the first 24 hours by treatment group. RIPC, remote ischaemic preconditioning; hs, high sensitivity; AUC diff, area under the curve difference.

**Figure 3.**
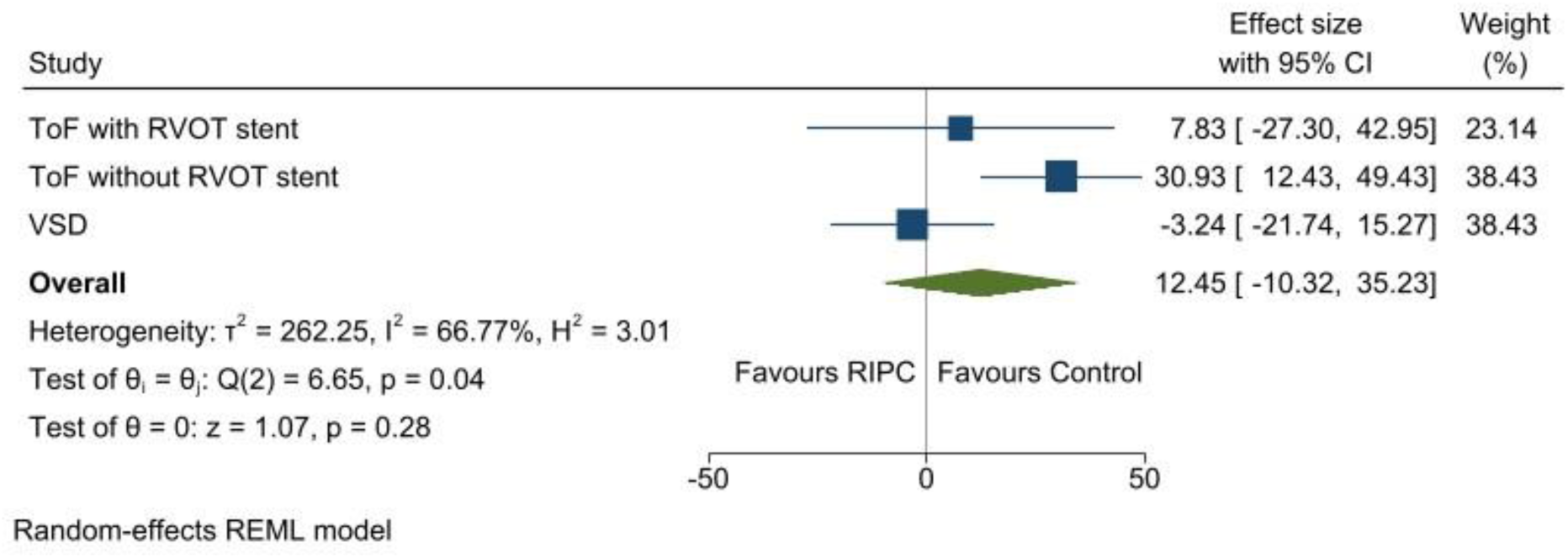
Forest plot of the primary outcome by congenital heart defect group. RIPC, remote ischaemic preconditioning; RVOT, right ventricular outflow tract; TOF, tetralogy of Fallot; VSD, ventricular septal defect.

**Table 3.** Mean plasma hs-troponin-T release at each time point, by treatment group.

| Time point | Plasma hs-troponin-T (µg/L), mean (SD, n) |  |
| --- | --- | --- |
|  | RIPC group | Control group |
| Baseline | 0.02 (0.02, 59) | 0.02 (0.02, 59) |
| 3 hours | 5.31 (3.66, 59) | 4.49 (2.56, 58) |
| 6 hours | 3.60 (2.65, 59) | 3.08 (1.68, 59) |
| 12 hours | 2.62 (1.85, 59) | 2.00 (1.02, 59) |
| 24 hours | 2.24 (2.19, 58) | 1.75 (1.23, 59) |

Postoperative time points from first release of the aortic cross-clamp. hs, high sensitivity; RIPC, remote ischaemic preconditioning.

**Table 4.** Predefined sub-group analyses for the primary outcome, area under the curve for plasma hs-troponin-T release in the first 24 hours after reperfusion.

|  |  | **RIPC**<br>**Mean (SD, n)** | **Control**<br>**Mean (SD, n)** | **Interaction**<br>**p-value** | **Mean difference**<br>**(95% CI) <sup>1</sup>** |
| --- | --- | --- | --- | --- | --- |
| Congenital heart defect | TOF with RVOT stent | 74.4 (23.1, 7) | 67.8 (41.7, 7) | 0.04 | 7.8 (-27.7, 43.4) |
|  | TOF without RVOT stent | 102.5 (57.5, 25) | 69.3 (25.3, 25) |  | 30.9 (12.2, 49.6) |
|  | VSD | 35.0 (13.5, 24) | 39.3 (23.1, 26) |  | -3.2 (-22.0, 15.5) |
|  |  | **RIPC**<br>**Mean (SD, n)** | **Control**<br>**Mean (SD, n)** | **Interaction**<br>**p-value** | **Mean difference**<br>**(95% CI) <sup>2</sup>** |
| Preoperative oxygen saturations | <90% (cyanotic) | 98.5 (58.6, 19) | 73.6 (34.1, 20) | 0.25 | 23.5 (2.0, 45.1) |
|  | ≥90% (acyanotic) | 55.4 (40.0, 37) | 46.2 (23.1, 38) |  | 7.8 (-7.9, 23.4) |
| Age | <1 year | 79.2 (55.6, 41) | 56.3 (26.5, 42) | 0.06 | 20.8 (6.1, 35.4) |
|  | ≥1 year | 44.9 (21.2, 15) | 54.0 (39.0, 16) |  | -6.7 (-30.6, 17.3) |

Mean difference (RIPC – Control) calculated using linear regression model, adjusting for either <sup>1</sup> baseline troponin and centre, or <sup>2</sup> baseline troponin, congenital heart defect and centre. A negative difference favours the RIPC group. RIPC, remote ischaemic preconditioning; RVOT, right ventricular outflow tract; TOF, tetralogy of Fallot; VSD, ventricular septal defect.

### Secondary outcomes and adverse events

There were no differences between groups in any of the secondary outcome measures: peak troponin, vasoactive inotrope score, arterial lactate, or central venous oxygen saturations in the first 12 hours, or length of postoperative stay in the PICU or hospital (figure 4 and table 5). No immediate limb complications were observed and there were no differences in adverse events following surgery (table 6). There were 11 serious adverse events in nine patients: four in the RIPC group and five in the control group. These comprised two post-cardiotomy ECLS and nine early surgical or catheter reinterventions, of which three were for residual lesions: one enlargement of right ventricular outflow tract, one surgical closure of residual ventricular septal defect, and one catheter closure of residual ventricular septal defect and stenting of pulmonary artery. Further details by group are provided in the Supplementary materials.

**Figure 4.**
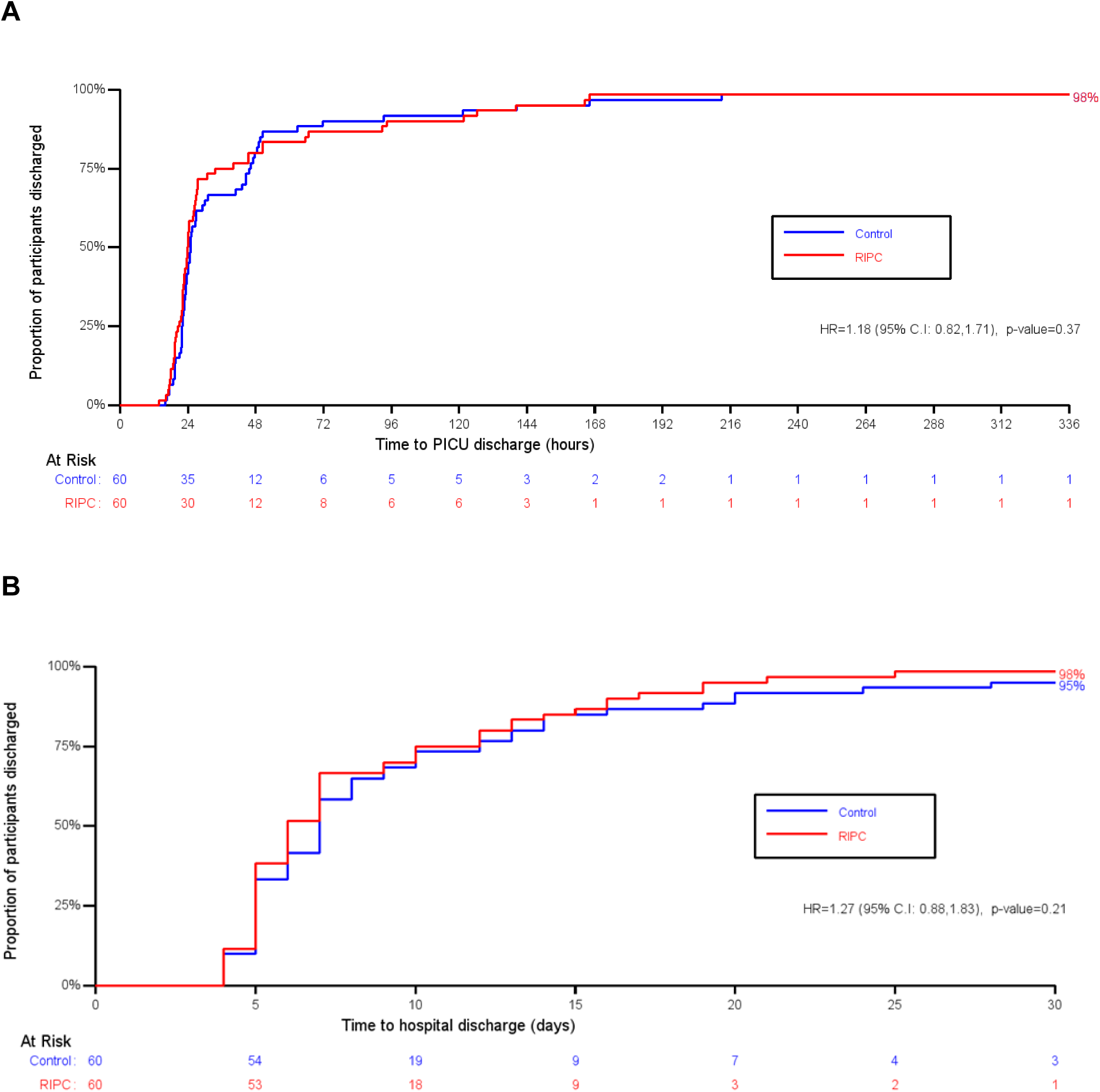
Kaplan Meier plots of time to discharge from A) paediatric intensive care unit, by group with data censored at 336 hours (14 days), and B) hospital, by treatment group with data censored at 30 days.

**Table 5.** Secondary outcomes by treatment group.

|  |  | **RIPC**<br>**Mean (SD, n)** | **Control**<br>**Mean (SD, n)** | **Point estimate**<br>**(95% CI), p value** |
| --- | --- | --- | --- | --- |
| Peak hs-troponin-T in first 12 hours (µg/L) |  | 5.42 (3.64, 57) | 4.51 (2.54, 58) | 0.85 (-0.09, 1.80), 0.08 <sup>1</sup> |
| Vasoactive inotrope score in first 12 hours |  | 61.6 (39.0, 60) | 53.6 (32.3, 60) | 8.1 (-4.1, 20.3), 0.19 <sup>2</sup> |
| Arterial lactate (mmol/L) | Baseline | 1.06 (0.29, 58) | 1.10 (0.56, 59) | 0.04 (-0.12, 0.20), 0.60 <sup>3</sup> |
|  | 3 hours | 1.69 (0.61, 60) | 1.61 (0.65, 59) |  |
|  | 6 hours | 1.73 (0.75, 57) | 1.73 (0.69, 58) |  |
|  | 9 hours | 1.50 (0.71, 55) | 1.62 (1.25, 55) |  |
|  | 12 hours | 1.45 (0.49, 55) | 1.47 (0.82, 55) |  |
| Central venous oxygen saturations (%) | 3 hours | 64.2 (10.2, 56) | 65.4 (8.7, 55) | -1.10 (-3.85, 1.66), 0.43 <sup>3</sup> |
|  | 6 hours | 61.9 (12.8, 55) | 63.9 (9.2, 57) |  |
|  | 9 hours | 60.7 (10.8, 50) | 62.2 (8.3, 50) |  |
|  | 12 hours | 61.4 (7.7, 51) | 61.8 (10.4, 57) |  |
|  |  | **RIPC**<br>**Median (IQR, n)** | **Control**<br>**Median (IQR, n)** | **Point estimate**<br>**(95% CI), p value <sup>4</sup>** |
| Time to PICU discharge (hours) |  | 23.92 (20.55, 33.58, 59) | 24.95 (22.00, 45.57, 59) | 1.18 (0.82, 1.71), 0.37 |
| Time to PICU discharge (hours) <sup>5</sup> |  | 23.96 (20.83, 36.78, 60) | 25.00 (22.02, 45.91, 60) | 1.20 (0.84, 1.73), 0.32 |
| Time to hospital discharge (days) |  | 6.0 (5.0, 10.0, 59) | 7.0 (5.0, 10.0, 57) | 1.27 (0.88, 1.83), 0.21 |
| Time to hospital discharge (days) <sup>5</sup> |  | 6.0 (5.0, 11.0, 60) | 7.0 (5.0, 12.0, 60) | 1.27 (0.88, 1.84), 0.20 |

Postoperative time points from first release of the aortic cross-clamp. Mean difference (RIPC – Control) calculated using linear regression model, adjusting for either <sup>1</sup> baseline troponin, congenital heart defect and centre, or <sup>2</sup> congenital heart defect and centre; a negative difference favours the RIPC group. <sup>3</sup> Average mean difference over each time point (RIPC – Control) calculated using a mixed linear regression model adjusting for baseline value of the measure (if available), heart condition and centre. For arterial lactate, a negative difference favours the RIPC group; for central venous oxygen saturations, a positive difference favours the RIPC group. <sup>4</sup> Hazard ratio (RIPC/Control) calculated using a Cox Proportional Hazards regression model, adjusted for centre and congenital heart defect; values greater than 1 favour RIPC. <sup>5</sup> Using uncensored data. PICU, paediatric intensive care unit; RIPC, remote ischaemic preconditioning.

**Table 6.** Adverse events and serious adverse events by treatment group.

|  | **RIPC (n=60)** | **Control (n=60)** |
| --- | --- | --- |
| Cardiac complication <sup>1</sup> | 7 (12%) | 6 (10%) |
| Cardiac arrest | 0 | 1 (2%) |
| ECLS required <sup>2</sup> | 1 (2%) | 1 (2%) |
| Post-operative complete heart block | 1 (2%) | 1 (2%) |
| Reoperation required <sup>2</sup> | 4 (7%) | 4 (7%) <sup>3</sup> |
| Pericardial collection/bleeding | 2 (3%) | 1 (2%) |
| Residual lesions | 1 (2%) | 2 (3%) |
| Epicardial pacemaker fitted | 1 (2%) | 1 (2%) |
| Chest left open | 4 (7%) | 2 (3%) |
| Post-CPB bleeding | 1 (2%) | 0 |
| Prolonged CPB time | 3 (5%) | 1 (2%) |
| Other | 0 | 1 (2%) |
| Neurological event <sup>2</sup> | 0 | 0 |
| Renal complication | 1 (2%) | 2 (3%) |
| Peak creatinine, µmol/L | 25 [20-31] | 24 [21-30] |
| Renal support, CVVH or PD | 1 (2%) | 2 (3%) |
| Respiratory event | 2 (3%) | 2 (3%) |
| Re-intubated | 0 | 1 (2%) |
| Infection – systemic | 4 (7%) | 1 (2%) |
| Infection – surgical site | 1 (2%) | 0 (0%) |
| Limb complication | 0 | 0 |
| Other adverse event | 0 | 0 |
| Death after surgery <sup>2</sup> | 0 | 0 |

Data are median [IQR] when appropriate. <sup>1</sup> Two participants experienced three adverse events each, whilst a further two participants experienced two adverse events each. In total, 19 adverse events occurred in 13 participants. <sup>2</sup> Serious adverse event requiring expedited reporting. <sup>3</sup> One participant underwent two re-operations to fit a temporary then permanent epicardial pacemaker; in total, five reoperations occurred in four participants. CPB, Cardiopulmonary Bypass; CVVH, Continuous Veno-Venous Haemofiltration; ECLS, Extracorporeal Life Support; PD, Peritoneal Dialysis.

### Metabolic phenotyping

Right atrial biopsies were collected at the onset of ischaemia (median 4 minutes after aortic cross-clamping) and at the end of ischaemia (median 53 minutes after aortic cross-clamping), and analysis was performed in 40 patients, 20 in each group. With correction for multiple testing (q<0.05), we found that in both early and late ischaemia, there were no differences in myocardial tissue metabolites or enrichment of metabolic pathways between patients receiving RIPC or sham intervention.

## DISCUSSION

In this trial, we found that adequately delivered, perioperative, bilateral RIPC, compared with sham inflation-deflation cycles, did not reduce myocardial injury in children undergoing elective surgery for common congenital heart defects. Troponin release was slightly higher in patients randomised to RIPC; this effect was increased in the per protocol analysis and was mainly attributable to greater postoperative troponin release in children with unstented TOF. This is the first trial to directly compare outcomes of RIPC in children with cyanotic and acyanotic congenital heart defects undergoing surgery at a similar age [18] and our findings suggest that bilateral RIPC may exacerbate myocardial ischaemia-reperfusion injury in children with TOF. RIPC did not alter the intensity of perioperative inotropic support, length of intensive care unit or hospital stay, or postoperative complications. The immediate clinical consequence of our results is to avoid remote ischemic preconditioning during paediatric cardiac surgery.

Overall AUC hs-troponin-T was higher in patients with TOF compared with those with an isolated VSD (Supplementary figure S1), reflecting a longer ischaemic time and the more frequent need for RVOT muscle resection. On sub-group analysis, we found no difference in troponin release related to preoperative oxygen saturations above or below 90%, but there was some evidence of an interaction with age (p=0.06), suggestive of greater myocardial injury in those aged less than 1 year who received RIPC, independent of defect type.

The promise of this simple, low-risk, inexpensive and readily available intervention as an adjunct to current methods for myocardial protection during cardiac surgery has prompted numerous trials in adults [26–31] and children [7–15] but with mixed results. A meta-analysis suggested that RIPC reduces myocardial injury during cardiac surgery [32], but subsequently two large multi-centre trials in adults failed to show benefit in either composite cardiovascular endpoints or troponin release [30,31]; both were criticised for using propofol anaesthesia after it had been shown to interfere with the preconditioning pathway [16,17]. In a more recent meta-analysis in children, including 793 patients in 9 trials, Tan et al determined that RIPC has a cardioprotective effect, with reduced troponin release at 6 hours, lower inotrope scores in the first 12 hours, and reduced PICU stay following surgery [33]; however they were unable to include the only large trial in most analyses due to a lack of suitable published data. Our trial makes an important contribution to the literature, raising significant doubts about the use of RIPC in paediatric cardiac surgery.

Cheung et al first demonstrated reductions in troponin release and perioperative inotropic requirements with RIPC in a heterogeneous cohort of 37 children, most of whom had either TOF or VSD, a similar population to our trial [7]. Several small studies found improved myocardial protection, with reduced early troponin release in infants and young children undergoing VSD closure [8,9], whilst others found no benefit in neonates or infants undergoing other operations for congenital heart disease [10-12,15]. In the largest paediatric trial of RIPC to date, McCrindle et al found no benefit in clinical outcomes, physiological biomarkers, or subgroup analyses in a mixed cohort of 299 children aged 0-17 years in Toronto, Canada [13] and proposed that better than expected outcomes in the control group, heterogeneity of underlying conditions, and use of propofol may have affected their findings. Failure to elicit a stimulus may also have been a key factor; manual inflation of the cuff to just 15mmHg above systolic pressure may have led to periods of subclinical reperfusion and the abolition of any preconditioning response. However, our findings support those of McCrindle et al that RIPC has little or no effect on clinical endpoints and therefore its role as a protective strategy in children undergoing surgery for congenital heart disease is limited.

A recent double-blind, randomised trial of 112 children undergoing complete repair of TOF in Wuhan, China found a reduced duration of PICU stay and lower AUC for troponin-T release in the first 24 hours with RIPC versus control [14]. Whilst patients underwent surgery at a similar age (11 months) to those in our trial, they were primarily acyanotic with mean preoperative oxygen saturations of 97-98% (SD ±3-4%), which is markedly different to our unstented TOF group (median 88%, IQR 86-97) in whom recent hypercyanotic spells were common. The authors also did not mention any pre-repair interventions for severe cyanosis, either Blalock-Thomas-Taussig shunt or RVOT stenting. These factors suggest that the pattern of TOF in their population is different to that seen most commonly in Europe and North America [34], or elsewhere in China [35]. This may be due to the higher incidence of a fibrous rather than muscular outlet septum in TOF in parts of East Asia, reducing the extent of RVOT obstruction and cyanosis [36], and explain the divergence of findings.

The mechanism underlying RIPC and its potential interaction with chronic cyanosis in the ischaemia-reperfusion injury associated with cardioplegic arrest is not clearly understood. RIPC has been shown to induce regulatory phosphorylation of key intracellular proteins involved in pro-survival metabolic signalling pathways [37,38]. Pepe et al evaluated the effect of RIPC on phosphorylated protein signalling in cyanotic children with TOF undergoing surgical repair in Melbourne, Australia. They found that signalling pathways were upregulated in resected RVOT muscle from both groups, supporting the hypothesis that the children may already have been ‘preconditioned’ by exposure to chronic hypoxaemia since birth, thereby pre-empting any potential benefit from perioperative RIPC [11]. In the same cohort, Hepponstall et al reported plasma proteomic changes in the RIPC group in the early postoperative period, with higher expression of proteins involved in metabolism, haemostasis, immunity, and inflammation that may be involved in RIPC-mediated cellular protection [39]. In experimental models of ischaemia-reperfusion, accumulation of succinate is a key metabolic signature of ischaemia and drives the generation of mitochondrial reactive oxygen species during reperfusion, leading to cellular injury [40]. We therefore postulated that RIPC may impact on the metabolic phenotype of the myocardium during ischaemia, but our analysis of right atrial samples taken in early and late ischaemia found no significant differences in metabolites or metabolic pathways between the RIPC and control arms. These novel findings suggest that downstream myocardial metabolism does not play a major role in mediating the effects of RIPC on the myocardium in children undergoing surgery with cardioplegic arrest.

In this trial, we addressed the methodological concerns raised over previous trials in children. A pressure-controlled digital tourniquet system was used, set to at least 50mmHg above systolic pressure to avoid sub-clinical reperfusion during the ischaemic phase of each cycle [13], with loss of arterial flow confirmed by distal pulse oximetry. A more intensive two cuff technique was used [29], applying a concurrent stimulus to both lower limbs to compensate for the lower skeletal muscle mass in young children. We avoided propofol anaesthesia which has been shown to attenuate the effects of RIPC [16,17,23] and excluded infants aged less than three months in whom the immature myocardium may be less responsive to RIPC [12]. Finally, we only sought to exploit the first window of preconditioning, performing the intervention under general anaesthesia prior to sternotomy, to avoid the potential distress, logistical challenges, and risks of incomplete intervention or withdrawal if applying RIPC to awake children at least 12 hours prior to surgery to exploit the second window of preconditioning [41]. This trial is the first multi-centre cardiac surgical trial in children in the UK [42] and demonstrates the value of collaboration to achieve recruitment targets in this challenging field.

### Strengths and limitations

The randomised, double-blind design, robust and verified delivery of bilateral RIPC, and quantification of a validated biomarker outcome are key strengths of our study. The results are consistent across analyses, including sensitivity analyses and comparison of secondary outcomes. The sample size calculation was based on analysis of the primary outcome in all participants and therefore sub-group analyses by the type of congenital heart defect, presence of cyanosis, and age should be seen as exploratory. It was also based on published studies using standard (fourth-generation) troponin assays, whereas we used a high-sensitivity (fifth-generation) assay which is now the preferred biomarker for determining myocardial injury [43]. Whilst troponin isoforms may be released from injured skeletal muscle [44], we found no difference between the RIPC and control groups in patients with a VSD, reassuring that the bilateral RIPC stimulus did not cause significant peripheral troponin-T release. Resection of hypertrophic septoparietal trabeculae from the right ventricular infundibulum and/or removal of an RVOT stent is likely to have increased troponin release, narrowing the effect of RIPC and predisposing to a type II error; despite this, RIPC remained associated with a greater AUC for troponin release in patients with TOF. We defined preoperative cyanosis as resting oxygen saturations in room air of <90% but TOF is a dynamic condition in which the right-to-left shunt and saturations may fluctuate depending on the clinical status of the child, weakening the use of a single measurement to determine dichotomous groups; this may have partly accounted for the differential effects of RIPC seen by congenital heart defect versus by preoperative oxygen saturations. The metabolomics analyses, while supporting the main finding, were performed in biopsies taken during early and late ischaemia which may not reflect any metabolic effects of RIPC on the baseline, pre-ischaemic myocardium.

Children undergoing surgery with cardioplegic arrest are at risk of perioperative myocardial injury, especially those with preoperative cyanosis [5,6]. Minimising damage to organs during heart surgery to reduce the frequency and severity of complications was recently identified as the #1 priority for research in children with congenital heart disease [45]. Future studies should focus on improving myocardial protection through multi-centre trials of established techniques, e.g. comparing different cardioplegia solutions, better understanding the mechanisms affecting ischaemic tolerance, including the impact of age and cyanosis, and moving towards personalised care through the application of genomic technology to explore phenotype-genotype interactions.

### Conclusions

Bilateral remote ischemic preconditioning does not improve cardioprotection in young children undergoing operative repair of common congenital heart defects and may be harmful in those with unstented TOF. The routine use of RIPC therefore cannot be recommended in paediatric cardiac surgery and alternative methods to improve myocardial protection and outcomes from surgery for congenital heart disease should be sought.

## Data Availability

Requests for access to data should be addressed to the Chief Investigator. Individual participant data collected during the trial (including the data dictionary) will be available, after deidentification, when the article has been published with no end date. All proposals requesting data access must specify how the data will be used, and all proposals will need the approval of the Trial Management Committee before data release.

## Funding

British Heart Foundation Intermediate Clinical Research Fellowship (grant number FS/15/49/31612) to NED; Medical Research Council UK (grant number MR/M009157/1) to Phenome Centre Birmingham.

## Acknowledgements

We thank Dr John V Pappachan (Southampton, UK) for guidance on the delivery of RIPC, Prof Andrew N Redington (Cincinnati, OH) for advice on RIPC and the exclusion criteria, Dr Oliver Stumper (Birmingham, UK) for his advice on ICON monitoring, and Prof Peter Brocklehurst, former Director of Birmingham Clinical Trials Unit, for his support for the trial. We are grateful to Monica Mitchell, Maureen Kelly, Kate Hong, George Preece, Helen Winmill and the PICU research nursing team for helping to conduct the trial. We thank Louise Dain, Louise Carlton, Lynda O’Connor, and Emily Scriven for their assistance in delivering the intervention. We thank laboratory staff at Birmingham Children’s Hospital: Fionnuala Terry, Anas Ghivalla, Ian Surplice, Siobhan McLean, Victoria Hendry, Elizabeth Bailey, Susan Betham, at Leeds Children’s Hospital: Jodie Sedgwick, Helena Baker, at Black Country Pathology Services: Dr Pervaz Mohammed, Dr Jonathan Berg, Rajvinder Garcha-Dadrah, Nirav Patel, Leena Kaur Bhachu, James Pethick, Waqqas Usmani, and at Phenome Centre Birmingham: Dr Catherine Winder. We are most grateful to Martina Ponsonby and the trustees of *Young at Heart* for their feedback on the study documents.

## Trial Management Committee

Mr Nigel E Drury (Chief Investigator), Mr Timothy J Jones (PI Birmingham), Ms Carin van Doorn (PI Leeds), Prof Paulus Kirchhof, Rehana Bi, Alicia Gill, Natalie J Ives, Dr Melanie Madhani, Dr James Montgomerie, Prof Kevin P Morris, Collette Spencer, John Stickley, Rebecca L Woolley.

## Data Monitoring Committee

Prof Gavin J Murphy (Chair), University of Leicester, UK; Dr Katherine L Brown, Great Ormond Street Hospital, London, UK; and Dr Peter Nightingale (Statistician), Queen Elizabeth Hospital Birmingham, UK.

## Clinical teams

Surgeons – Birmingham: Mr Timothy Jones, Prof David Barron, Mr Phil Botha, Ms Natasha Khan, and Leeds: Ms Carin van Doorn, Mr Osama Jaber, Mr Imre Kassai, Mr Guiseppe Pelella. Anaesthetists – Birmingham: Dr James Montgomerie, Dr Edmund Carver, Dr Alistair Cranston, Dr Fraser Harban, Dr Vasco Laginha Rolo, Dr Ritchie Marcus, Dr Tony Moriarty, Dr Raju Reddy, Dr Susanna Ritchie-McLean, Dr Monica Stokes, Dr Ayngara Thillaivasan, and Leeds: Dr Nandlal Bhatia, Dr Carol Bodlani, Dr Wendy Lim, Dr Joe Mellor, Dr Jutta Scheffczik.

## Conflicts of Interest

None declared.

## Notes

### Competing Interest Statement

The authors have declared no competing interest.

### Clinical Trial

ISRCTN12923441

### Funding Statement

The study was funded by the British Heart Foundation through an Intermediate Clinical Research Fellowship (FS/15/49/31612) awarded to Nigel Drury. Phenome Centre Birmingham was supported by a grant from the Medical Research Council UK (MR/M009157/1).

### Author Declarations

The West Midlands-Solihull National Health Service Research Ethics Committee gave ethical approval for this study on 5 August 2016 (ref: 16/WM/0309).

