## Supplementary materials for "The Bilateral Remote Ischaemic Conditioning in Children (BRICC) trial: a two-centre, double-blind, randomised controlled trial in young children undergoing cardiac surgery"

### Supplementary Methods

Only patients with the most common form of tetralogy of Fallot were included; variants such as absent pulmonary valve syndrome, pulmonary atresia with major aortopulmonary collateral arteries, or with an atrioventricular septal defect were not included. Regulatory approval from the Medicines and Healthcare products Regulatory Agency (MHRA) was not required as this trial was not a clinical trial of an investigational medicinal product (CTIMP).

*Cardiac output measurement*

Cardiac output was measured as an exploratory endpoint using ICON (Osypka Medical, Berlin), a CE-marked, FDA-approved device that uses non-invasive electrical velocimetry to provide a continuous assessment of haemodynamic parameters that has been validated in infants in the early postoperative period [1,2]. Cardiac index was determined during the first 12 hours after reperfusion in patients undergoing surgery in Birmingham only and analysed using mixed effect repeated measures models using all available data, presented as an adjusted mean difference and 95% confidence interval.

*Metabolic phenotyping*

Untargeted metabolomic analysis was performed applied. Tissue samples were extracted applying a biphasic extraction protocol using chloroform ($CHCl_3$), methanol (MeOH) and water ($H_2O$) and dried as was previously described [3]. Four complementary ultra high performance liquid chromatography-mass spectrometry assays were applied to analyse each sample extract, as previously described, including the analysis of pooled QC samples and process blank samples [4]. Vendor format raw data files (.RAW) were converted to the mzML file format using ProteoWizard software [4]. Deconvolution was performed by XCMS software 4 (version 1.46.0 running in the Galaxy environment) [4]. QC samples with a total peak area (of all features) exceeding ±25% of the median QC total peak area were removed from the data matrix. Features were retained in the data matrix if they were: present in >90% of QC samples; had a peak area relative standard deviation (RSD) <30% across QC samples; and had an extract blank/mean QC area ratio of <5%. Features with a <50% detection rate over all samples were also removed. Univariate analysis was performed applying one-way ANOVA on PQN normalised and glog transformed data with a critical p-value >0.05 applied after correction for multiple testing applying the Benjamini-Hochberg procedure [5].

### Supplementary Results

The first patient was randomised on 24 October 2016 and the last patient completed follow-up on 21 December 2020.

*Covid-19 pandemic*

Along with all other non-Covid public health studies, recruitment to the trial was suspended on 13 March 2020 due to the impact of the Covid-19 pandemic on the National Health Service. During the suspension, 29 otherwise eligible patients underwent surgery at the two sites but had to be excluded from the trial. Recruitment recommenced on 29 June 2020 and all patients recruited subsequently were shown to be negative for SARS-CoV-2 with reverse-transcriptase polymerase chain reaction (PCR) on two consecutive swabs prior to undergoing surgery.

*Application of the intervention*

In the RIPC arm, the intervention was applied to both thighs in 55 (92%) patients and to one thigh and one upper arm in 5 (8%) patients due to one lower limb being unavailable. The mean limb occlusion pressures for the three cycles were 152 mmHg (+/-11), 156 mmHg (+/-18) and 156 mmHg (+/-12), respectively. Adherence to treatment allocation was achieved in 116 (96.7%) patients; no patients in the control arm received RIPC but 4 (7%) patients in the RIPC arm did not have three confirmed cycles of limb ischaemia-reperfusion, with continual loss of arterial flow confirmed by distal pulse oximetry during each period of limb ischaemia. There were no immediate or delayed limb complications due to the use of RIPC.

*Cardiac output measurement*

Haemodynamic assessment was performed using ICON in 82 children undergoing surgery in Birmingham only. There was no difference in cardiac index in the first 12 hours between the two groups, as shown in table S3. There was incomplete data recording in the postoperative period due to repeated electrode detachment or persistent movement artefact after waking, limiting its value as a routine monitoring tool in the setting of post-surgical ICU care.

*Serious adverse events*

There were 11 serious adverse events in nine patients: four in the RIPC group and five in the control group. These comprised two post-cardiotomy ECLS and nine early surgical or catheter reinterventions. All were assessed to be unrelated to the trial intervention and no actions were required by the sponsor, as shown in table S4.

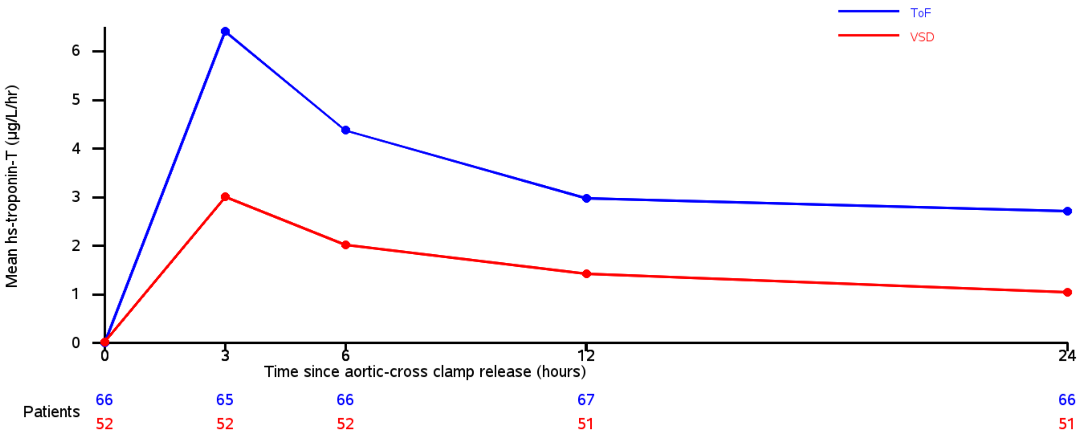


**Figure S1**. Mean hs-troponin-T release in the first 24 hours by congenital heart defect.

TOF, tetralogy of Fallot; VSD, ventricular septal defect.

**Table S1**. Preoperative oxygen saturations by congenital heart defect group.

| Preoperative oxygen saturations | Congenital heart defect | | |
|---|---|---|---|
| | TOF without RVOT stent (n=54) | TOF with RVOT stent (n=14) | VSD (n=52) |
| <90% (cyanotic) | 30 (56%) | 10 (71%) | 1 (2%) |
| ≥90% (acyanotic) | 24 (44%) | 4 (29%) | 51 (98%) |
| Median (IQR), % | 88 [86-97] | 83.5 [81-90] | 98 [97-99.5] |

RVOT, right ventricular outflow tract; TOF, tetralogy of Fallot; VSD, ventricular septal defect.

**Table S2**. Age category by congenital heart defect group.

| Age | Congenital heart defect | | |
|---|---|---|---|
| | TOF without RVOT stent (n=54) | TOF with RVOT stent (n=14) | VSD (n=52) |
| <1 year | 45 (83%) | 8 (57%) | 35 (67%) |
| ≥1 year | 9 (17%) | 6 (43%) | 17 (33%) |
| Median (IQR), months | 8.0 [5.0-10.0] | 10.5 [7.0-13.0] | 5.5 [3.5-14.5] |

RVOT, right ventricular outflow tract; TOF, tetralogy of Fallot; VSD, ventricular septal defect.

**Table S3**. Cardiac index in the first 12 hours measured using ICON, in Birmingham only.

| Cardiac index ($L/min/m^2$) | RIPC Mean (SD, n) | Control Mean (SD, n) | Point estimate (95% CI), p value [1] |
|---|---|---|---|
| Baseline | 3.7 (1.3, 41) | 4.1 (1.3, 41) | 0.22 (-0.16, 0.59) |
| 3 hours | 3.8 (1.4, 36) | 3.7 (1.1, 38) | |
| 6 hours | 3.7 (1.3, 39) | 3.8 (1.4, 37) | |
| 9 hours | 3.6 (1.0, 37) | 3.8 (1.4, 32) | |
| 12 hours | 3.6 (1.0, 35) | 3.8 (1.5, 29) | |

[1] Average mean difference over each time point (RIPC – Control) calculated using a mixed linear regression model adjusting for baseline value. A positive difference favours the RIPC group. RIPC, remote ischaemic preconditioning.

**Table S4**. Details of serious adverse events by group.

| | Event | Date | Description | Causality assessment | Action required |
|---|---|---|---|---|---|
| **RIPC** | | | | | |
| 1 | Reoperation required | 12/2016 | Severe tachycardia with poor ventricular function, pericardial collection, worsening acidosis, chest re-explored and left open. | Unrelated | None |
| 2 | ECLS required | 04/2019 | Severe low cardiac output following surgery requiring mechanical support with ECLS, chest already open post-op. | Unrelated | None |
| 2 | Reoperation required | 04/2019 | Following surgery, large residual shunt around VSD patch. Reoperation to close the residual defect. | Unrelated | None |
| 3 | Reoperation required | 05/2019 | Excessive bleeding via chest drains on PICU in early post op period. Return to theatre for reoperation; bleeding stopped. | Unrelated | None |
| 4 | Reoperation required | 08/2019 | Intermittent complete heart block following surgery requiring reoperation for epicardial pacemaker. | Unrelated | None |
| **Control** | | | | | |
| 5 | ECLS required | 10/2016 | Cardiac arrest on PICU during early post-operative period, requiring ECPR and mechanical support with ECLS. | Unrelated | None |
| 6 | Reoperation required | 08/2017 | Reoperation two weeks after repair of TOF, to enlarge the RVOT due to an increasing gradient on echo. | Unrelated | None |
| 7 | Reoperation required | 02/2018 | Re exploration for hypotension despite increasing inotropes, poor ventricular function, chest reopened and left open. | Unrelated | None |
| 8 | Reoperation required | 06/2018 | Residual shunt around VSD patch. Catheter procedure for VSD closure and stenting of the left pulmonary artery. | Unrelated | None |
| 9 | Reoperation required | 11/2019 | Post-op 2nd degree heart block, progressed to complete heart block, requiring insertion of epicardial pacing wire. | Unrelated | None |
| 9 | Reoperation required | 12/2019 | Persistent complete heart block and pacing dependent so redo-sternotomy for insertion of permanent epicardial system. | Unrelated | None |

ECLS, extracorporeal life support; ECPR, extracorporeal cardiopulmonary resuscitation; PICU, paediatric intensive care unit; RIPC, remote ischaemic preconditioning; RVOT, right ventricular outflow tract; TOF, tetralogy of Fallot; VSD, ventricular septal defect.
